# Screening of healthcare workers for SARS-CoV-2 highlights the role of asymptomatic carriage in COVID-19 transmission

**DOI:** 10.1101/2020.05.09.20082909

**Authors:** Lucy Rivett, Sushmita Sridhar, Dominic Sparkes, Matthew Routledge, Nick K. Jones, Sally Forrest, Jamie Young, Joana Pereira-Dias, William L. Hamilton, Mark Ferris, M. Estee Torok, Luke Meredith, The CITIID-NIHR COVID-19 BioResource Collaboration, Martin Curran, Stewart Fuller, Afzal Chaudhry, Ashley Shaw, Richard J. Samworth, John R. Bradley, Gordon Dougan, Kenneth G.C. Smith, Paul J. Lehner, Nicholas J. Matheson, Giles Wright, Ian Goodfellow, Stephen Baker, Michael P. Weekes

## Abstract

Significant differences exist in the availability of healthcare worker (HCW) SARS-CoV-2 testing between countries, and existing programmes focus on screening symptomatic rather than asymptomatic staff. Over a 3-week period (April 2020), 1,032 asymptomatic HCWs were screened for SARS-CoV-2 in a large UK teaching hospital. Symptomatic staff and symptomatic household contacts were additionally tested. Real-time RT-PCR was used to detect viral RNA from a throat+nose self-swab.

3% of HCWs in the *asymptomatic screening group* tested positive for SARS-CoV-2. 17/30 (57%) were truly asymptomatic/pauci-symptomatic. 12/30 (40%) had experienced symptoms compatible with coronavirus disease 2019 (COVID-19) >7 days prior to testing, most self-isolating, returning well. Clusters of HCW infection were discovered on two independent wards. Viral genome sequencing showed that the majority of HCWs had the dominant lineage B·1. Our data demonstrates the utility of comprehensive screening of HCWs with minimal or no symptoms. This approach will be critical for protecting patients and hospital staff.

**Appendix: The CITIID-NIHR COVID-19 BioResource Collaboration:** *Principal Investigators:* Stephen Baker, John Bradley, Gordon Dougan, Ian Goodfellow, Ravi Gupta, Paul J. Lehner, Paul A. Lyons, Nicholas J. Matheson, Kenneth G.C. Smith, M. Estee Torok, Mark Toshner, Michael P. Weekes

*Infectious Diseases Department:* Nicholas K. Jones, Lucy Rivett, Matthew Routledge, Dominic Sparkes, Ben Warne

*SARS-CoV-2 testing team:* Josefin Bartholdson Scott, Claire Cormie, Sally Forrest, Harmeet Gill, Iain Kean, Mailis Maes, Joana Pereira-Dias, Nicola Reynolds, Sushmita Sridhar, Michelle Wantoch, Jamie Young

*COG-UK Cambridge Sequencing Team:* Sarah Caddy, Laura Caller, Theresa Feltwell, Grant Hall, William Hamilton, Myra Hosmillo, Charlotte Houldcroft, Aminu Jahun, Fahad Khokhar, Luke Meredith, Anna Yakovleva

*NIHR BioResource:* Helen Butcher, Daniela Caputo, Debra Clapham-Riley, Helen Dolling, Anita Furlong, Barbara Graves, Emma Le Gresley, Nathalie Kingston, Sofia Papadia, Hannah Stark, Kathleen E. Stirrups, Jennifer Webster

*Research nurses:* Joanna Calder, Julie Harris, Sarah Hewitt, Jane Kennet, Anne Meadows, Rebecca Rastall, Criona O,Brien, Jo Price, Cherry Publico, Jane Rowlands, Valentina Ruffolo, Hugo Tordesillas

*NIHR Cambridge Clinical Research Facility:* Karen Brookes, Laura Canna, Isabel Cruz, Katie Dempsey, Anne Elmer, Naidine Escoffery, Stewart Fuller, Heather Jones, Carla Ribeiro, Caroline Saunders, Angela Wright

*Cambridge Cancer Trial Centre:* Rutendo Nyagumbo, Anne Roberts

*Clinical Research Network Eastern:* Ashlea Bucke, Simone Hargreaves, Danielle Johnson, Aileen Narcorda, Debbie Read, Christian Sparke, Lucy Warboys

*Administrative staff, CUH:* Kirsty Lagadu, Lenette Mactavous

*CUH NHS Foundation Trust:* Tim Gould, Tim Raine, Ashley Shaw

*Cambridge Cancer Trials Centre:* Claire Mather, Nicola Ramenatte, Anne-Laure Vallier

*Legal/Ethics:* Mary Kasanicki

*CUH Improvement and Transformation Team:* Penelope-Jane Eames, Chris McNicholas, Lisa Thake Clinical Microbiology & Public Health Laboratory (PHE): Neil Bartholomew, Nick Brown, Martin Curran, Surendra Parmar, Hongyi Zhang

*Occupational Health:* Ailsa Bowring, Mark Ferris, Geraldine Martell, Natalie Quinnell, Giles Wright, Jo Wright

*Health and Safety:* Helen Murphy

*Department of Medicine Sample Logistics:* Benjamin J. Dunmore, Ekaterina Legchenko, Stefan Gräf, Christopher Huang, Josh Hodgson, Kelvin Hunter, Jennifer Martin, Federica Mescia, Ciara O’Donnell, Linda Pointon, Joy Shih, Rachel Sutcliffe, Tobias Tilly, Zhen Tong, Carmen Treacy, Jennifer Wood Department of Medicine Sample Processing and Acquisition: Laura Bergamaschi, Ariana Betancourt, Georgie Bowyer, Aloka De Sa, Maddie Epping, Andrew Hinch, Oisin Huhn, Isobel Jarvis, Daniel Lewis, Joe Marsden, Simon McCallum, Francescsa Nice, Ommar Omarjee, Marianne Perera, Nika Romashova, Mateusz Strezlecki, Natalia Savoinykh Yarkoni, Lori Turner

*Epic team/other computing support:* Barrie Bailey, Afzal Chaudhry, Rachel Doughton, Chris Workman

*Statistics/modelling:* Richard J. Samworth, Caroline Trotter

## Introduction

Despite the World Health Organisation (WHO) advocating widespread testing for SARS-CoV-2, national capacities for implementation have diverged considerably.^1,2^ In the UK, the strategy has been to perform SARS-CoV-2 testing for essential workers who are symptomatic themselves or have symptomatic household contacts. This approach has been exemplified by recent studies of symptomatic HCWs.^3,4^ The role of nosocomial transmission of SARS-CoV-2 is becoming increasingly recognised, accounting for 12-29% of cases in some reports.^5^ Importantly, data suggest that the severity and mortality risk of nosocomial transmission may be greater than for community-acquired COVID-19.^6^

Protection of HCWs and their families from the acquisition of COVID-19 in hospitals is paramount, and underscored by rising numbers of HCW deaths nationally and internationally.^7,8^ In previous epidemics, HCW screening programmes have boosted morale, decreased absenteeism and potentially reduced long-term psychological sequelae.^9^ Screening also allows earlier return to work when individuals or their family members test negative.^3,4^ Another major consideration is the protection of vulnerable patients from a potentially infectious workforce^6^, particularly as social distancing is not possible whilst caring for patients. Early identification and isolation of infectious HCWs may help prevent onward transmission to patients and colleagues, and targeted infection prevention and control measures may reduce the risk of healthcare-associated outbreaks.

The clinical presentation of COVID-19 can include minimal or no symptoms.^10^ Asymptomatic or pre-symptomatic transmission is clearly reported and is estimated to account for around half of all cases of COVID-19.^11^ Screening approaches focussed solely on symptomatic HCWs are therefore unlikely to be adequate for suppression of nosocomial spread. Preliminary data suggests that mass screening and isolation of asymptomatic individuals can be an effective method for halting transmission in community-based settings.^12^ Recent modelling has suggested that weekly testing of asymptomatic HCWs could reduce onward transmission by 16-23%, on top of isolation based on symptoms, provided results are available within 24 hours.^13^ The need for widespread adoption of an expanded screening programme for asymptomatic, as well as symptomatic HCWs, is apparent.^13-15^

Challenges to the roll-out of an expanded screening programme include the ability to increase diagnostic testing capacity, logistical issues affecting sampling and turnaround times and concerns about workforce depletion should substantial numbers of staff test positive. Here, we describe how we have dealt with these challenges and present initial findings from a comprehensive staff screening programme at Cambridge University Hospitals NHS Foundation Trust (CUHNFT). This has included systematic screening of >1,000 asymptomatic HCWs in their workplace, in addition to >200 symptomatic staff or household contacts. Screening was performed using a validated real-time reverse transcription PCR (RT-PCR) assay detecting SARS-CoV-2 from combined oropharyngeal (OP) and nasopharyngeal (NP) swabs.^16^ Rapid viral sequencing of positive samples was used to further assess potential epidemiological linkage where nosocomial transmission was suspected. Our experience highlights the value of programmes targeting both symptomatic and asymptomatic staff, and will be informative for the establishment of similar programmes in the UK and globally.

## Results

### Characteristics of HCW and testing groups

Between 6^th^ and 24^th^ April 2020, 1,270 HCWs in CUHNFT and their symptomatic household contacts were swabbed and tested for SARS-CoV-2 by real-time RT-PCR. The median age of the HCWs was 34; 71% were female and 29% male. The technical RT-PCR failure rate was 2/1,270 (0·2% see Methods); these were excluded from the ‘Tested’ population for further analysis. Ultimately, 5% (n=61) of swabs were SARS-CoV-2 positive. 21 individuals underwent repeat testing for a variety of reasons, including evolving symptoms (n=3) and scoring ‘medium’ probability on clinical COVID-19 criteria (Tables S1-S2) (n=11). All remained SARS-CoV-2 negative. Turn around time from sample collection to resulting was 12-36 hours; this varied according to the time samples were obtained.

Table 1 outlines the total number of SARS-CoV-2 tests performed in each screening group (*HCW asymptomatic, HCW symptomatic*, and *HCW symptomatic household contact*) categorised according to the ward with the highest anticipated risk of exposure to COVID-19 (‘red’, high; ‘amber’, medium; ‘green’, low; table S1). In total, 31/1,032 (3%) of those tested in the *HCW asymptomatic screening group* tested SARS-CoV-2 positive. In comparison, 30/221 (14%) tested positive when *HCW symptomatic* and *HCW symptomatic household contact screening groups* were combined. As expected, symptomatic HCWs and their household contacts were significantly more likely to test positive than HCWs from the *asymptomatic screening group* (p<0·0001, Fisher’s exact test). HCWs working in ‘red’ or ‘amber’ wards were significantly more likely to test positive than those working in ‘green’ wards (p=0·0042, Fisher’s exact test).

**Table 1.** Total number of SARS-CoV-2 tests performed in each screening group categorised according to the highest risk ward of potential exposure.

| Clinical Area |  |  |  |  |  |
| --- | --- | --- | --- | --- | --- |
|  | Green | Amber | Red | Unknown | Total |
| *HCW asymptomatic* | 7/454 | 4/78 | 20/466 | 0/34 | **31/1032** |
| *screening group* | (1.5%) | (5.1%) | (4.3%) | (0%) | **(3%)** |
| *HCW symptomatic* | 8/66 | 1/9 | 17/88 | 0/6 | **26/169** |
| *screening group* | (12.1%) | (11.1%) | (19.3%) | (0%) | **(15.4%)** |
| *HCW symptomatic* | 2/14 | 0/1 | 0/14 | 2/23 | **4/52** |
| *household contacts* | (14.3%) | (0%) | (0%) | (8.7%) | **(7.7%)** |
| *Unknown* | 0/4 (0%) | 0/0 | 0/7 (0%) | 0/4 (0%) | **0/15 (0%)** |
| *All* | **17/538** | **5/88** | **37/575** | **2/67** | **61/1268** |
|  | **(3.2%)** | **(5.7%)** | **(6.4%)** | **(3%)** | **(4.8%)** |

Viral loads varied between individuals, potentially reflecting the nature of the sampling site. However, for individuals testing positive for SARS-CoV-2, viral loads were significantly lower for those in the *HCW asymptomatic screening group* than in those tested due to the presence of symptoms (Figure 1). For the *HCW symptomatic* and *HCW symptomatic contact screening groups*, viral loads did not correlate with duration of symptoms or with clinical criteria risk score (Figure S1 and data not shown).

**Figure 1.**
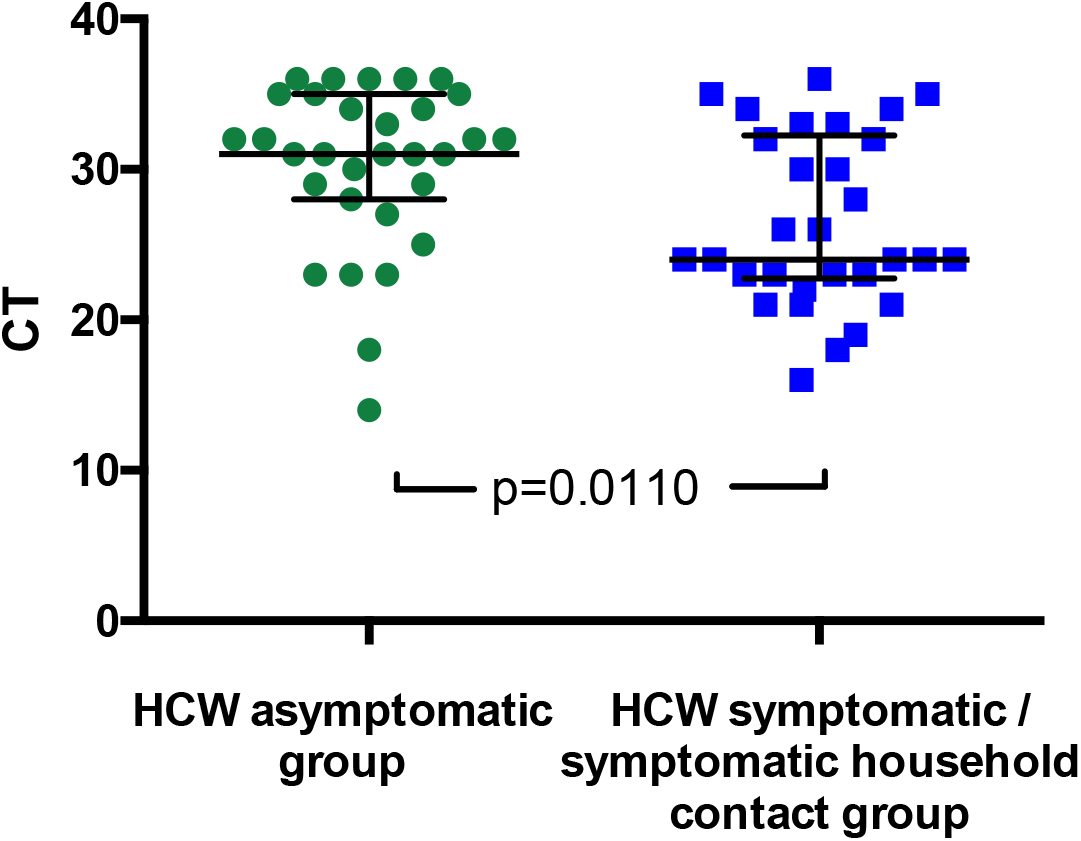
SARS-CoV-2 RNA CT (cycle threshold) values for those individuals who tested positive shown according to HCW group: *HCW asymptomatic screening group* (green circles); *HCW symptomatic* or *symptomatic household contact screening groups* (blue squares). A Mann Whitney test was used to compare the two groups. Bars: median +/- interquartile range. Note that lower CT values correspond to earlier detection of the viral RNA in the RT-PCR process and therefore identify swabs with higher numbers of copies of the viral genome.

### Three subgroups of SARS-CoV-2 positive asymptomatic HCW

Each individual in the *HCW asymptomatic screening group* was contacted by telephone to establish a clinical history, and COVID-19 probability criteria (Table S3) were retrospectively applied to categorise any symptoms in the month prior to testing (Figure 2). One HCW could not be contacted to obtain further history. Individuals captured by the *HCW asymptomatic screening group* were generally *asymptomatic at the time of screening*, however could be divided into three sub-groups: (i) HCWs with no symptoms at all, (ii) HCWs with (chiefly low-to-medium COVID-19 probability) symptoms commencing ≤7 days prior to screening and (iii) HCWs with (typically high COVID-19 probability) symptoms commencing >7 days prior to screening (Figure 2). 9/12 (75%) individuals with symptom onset >7 days previously had appropriately self-isolated and then returned to work. One individual with no symptoms at the time of swabbing subsequently developed symptoms prior to being contacted with their positive result. Overall, 5/1032 (0.5%) individuals in the asymptomatic screening group were identified as truly asymptomatic carriers of SARS-CoV-2, and 1/1032 (0.1%) was identified as pre-symptomatic. Table 2 shows illustrative clinical vignettes.

**Figure 2:**
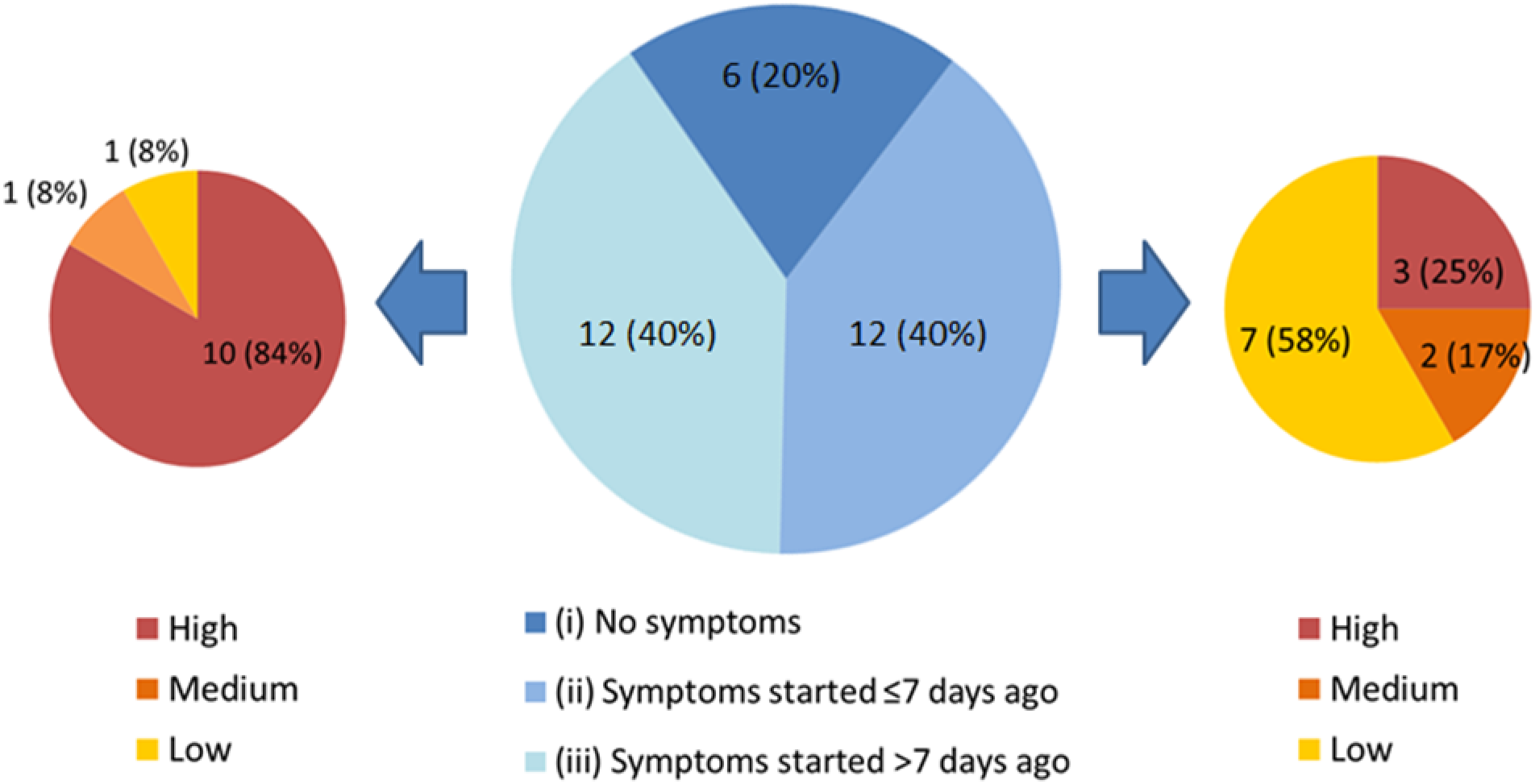
Three subgroups of staff testing SARS-CoV-2 positive from the *HCW asymptomatic screening group* (central pie chart, described in detail in the main text). n=number of individuals (% percentage of total). The peripheral pie charts show number and percentage of individuals in groups (ii – right pie chart) and (iii – left pie chart) with low, medium and high COVID-19 probability symptoms upon retrospective analysis.

**Table 2:**
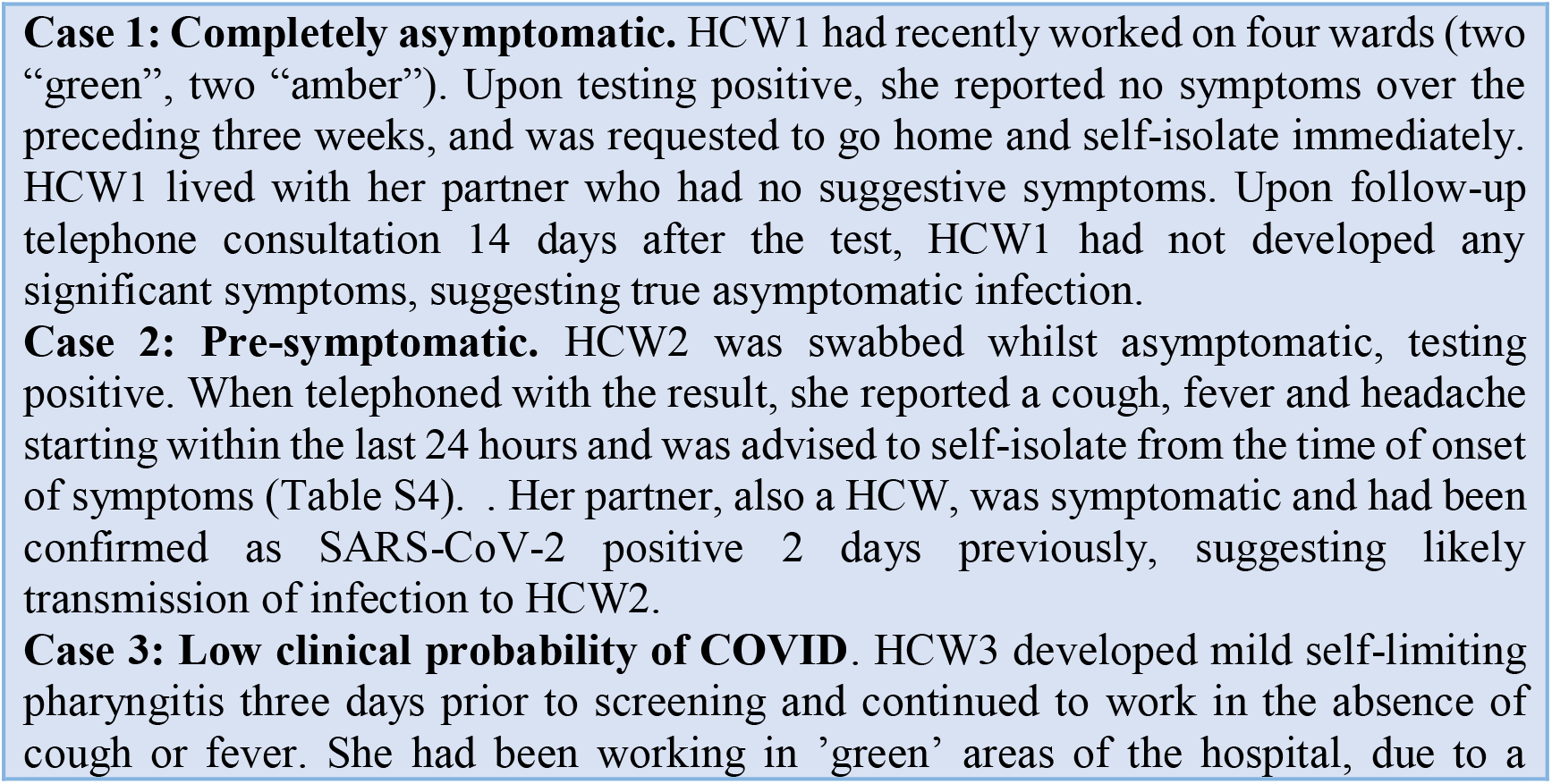
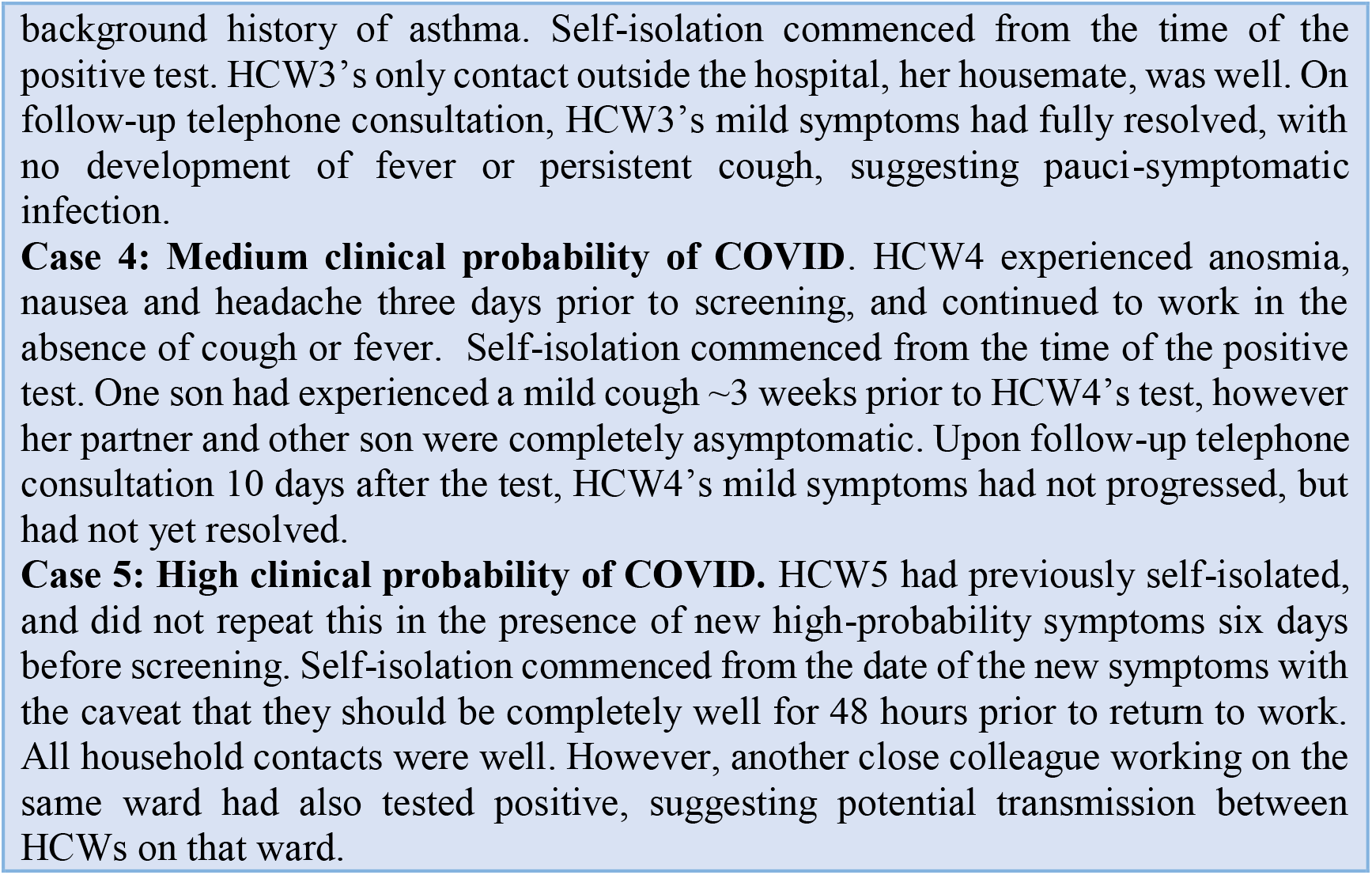
Clinical vignettes. Self-isolation instructions were as described in Table S4.

### Identification of two clusters of cases by screening asymptomatic HCWs

For the *HCW asymptomatic screening group*, nineteen wards were identified for systematic priority screening as part of hospital-wide surveillance. Two further areas were specifically targeted for screening due to unusually high staff sickness rates (ward F), or concerns about appropriate PPE usage (ward Q) (Figure 3, Table S5). Interestingly, in line with findings in the total HCW population, a significantly greater proportion of HCWs working on ‘red’ wards compared to HCWs working on ‘green’ wards tested positive as part of the asymptomatic screening programme (‘green’ 6/310 vs ‘red’ 19/372; p=0·0389, Fisher’s exact test). The proportion of HCW with a positive test was significantly higher on Ward F than on other wards categorised as ‘green’ clinical areas (ward F 4/43 vs other ‘green’ wards 2/267; p=0·0040, Fisher’s exact test). Likewise, amongst wards in the ‘red’ areas, ward Q showed significantly higher rates of positive HCW test results (ward Q 7/37 vs other ‘red’ wards 12/335; p=0·0011, Fisher’s exact test).

**Figure 3:**
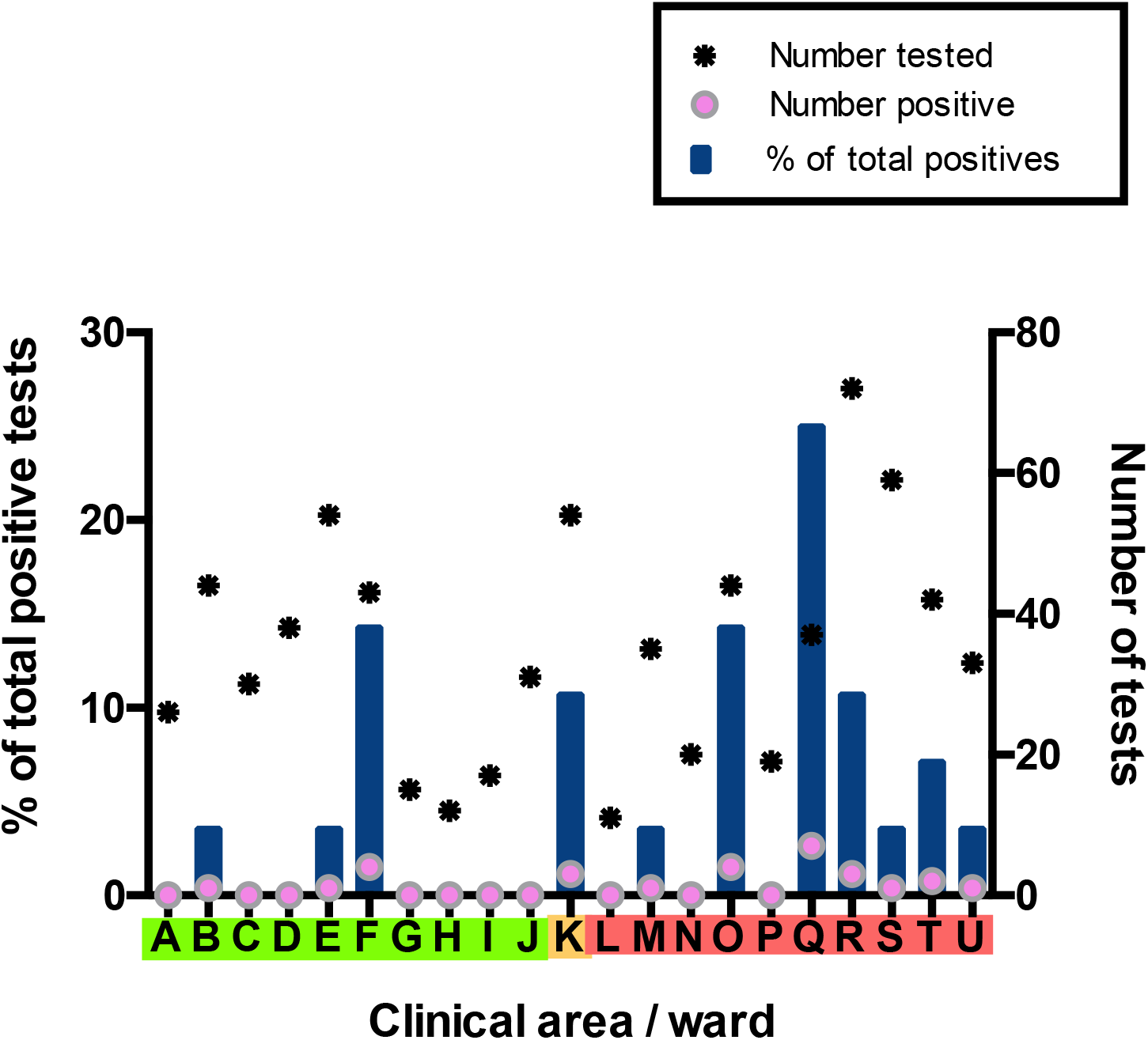
Distribution of SARS-CoV-2 positive cases across 21 clinical areas, detected by ward-based asymptomatic screening (underlying data shown in Table S5). Wards are coloured (‘green’, ‘amber’, ‘red’) according to risk of anticipated exposure to COVID-19 (Table S1). HCWs working across >1 ward were counted for each area. The left-hand y-axis shows the percentage of positive results from a given ward compared to the total positive results from the *HCW asymptomatic screening group* (blue bars). The right-hand y-axis shows the total number of SARS-CoV-2 tests (stars) and the number positive (pink circles). Asymptomatic screening tests were also performed on an opportunistic basis in individuals working outside these areas, as well as in a subsequent intensified manner on ward F and ward Q after identification of clusters of positive cases on these wards (Figure 4). Results of these tests are included in summary totals in Table 1, but not in this figure.

Ward F is an elderly care ward, designated as a ‘green’ area with Scenario 0 PPE (Tables S3-S4), with a high proportion of COVID-19 vulnerable patients due to age and comorbidity. 4/43 (9%) ward staff tested positive for SARS-CoV-2. In addition, two staff members on this ward tested positive in the *HCW symptomatic/symptomatic contact screening groups*. All positive HCWs were requested to self-isolate, the ward was closed to admissions and escalated to Scenario 1 PPE (Table S2). Reactive screening of a further 18 ward F staff identified an additional three positive asymptomatic HCWs (Figure 4). Sequence analysis indicated that 6/9 samples from HCW who worked on ward F belonged to SARS-CoV-2 lineage B·1 (currently known to be circulating in at least 43 countries^20^), with a further two that belonged to B1·7 and one that belonged to B2·1. This suggests more than two introductions of SARS-CoV-2 into the HCW population on ward F (Figures S2-S3, Table S6). It was subsequently found that two further staff members from ward F had previously been admitted to hospital with severe COVID-19 infection.

**Figure 4.**
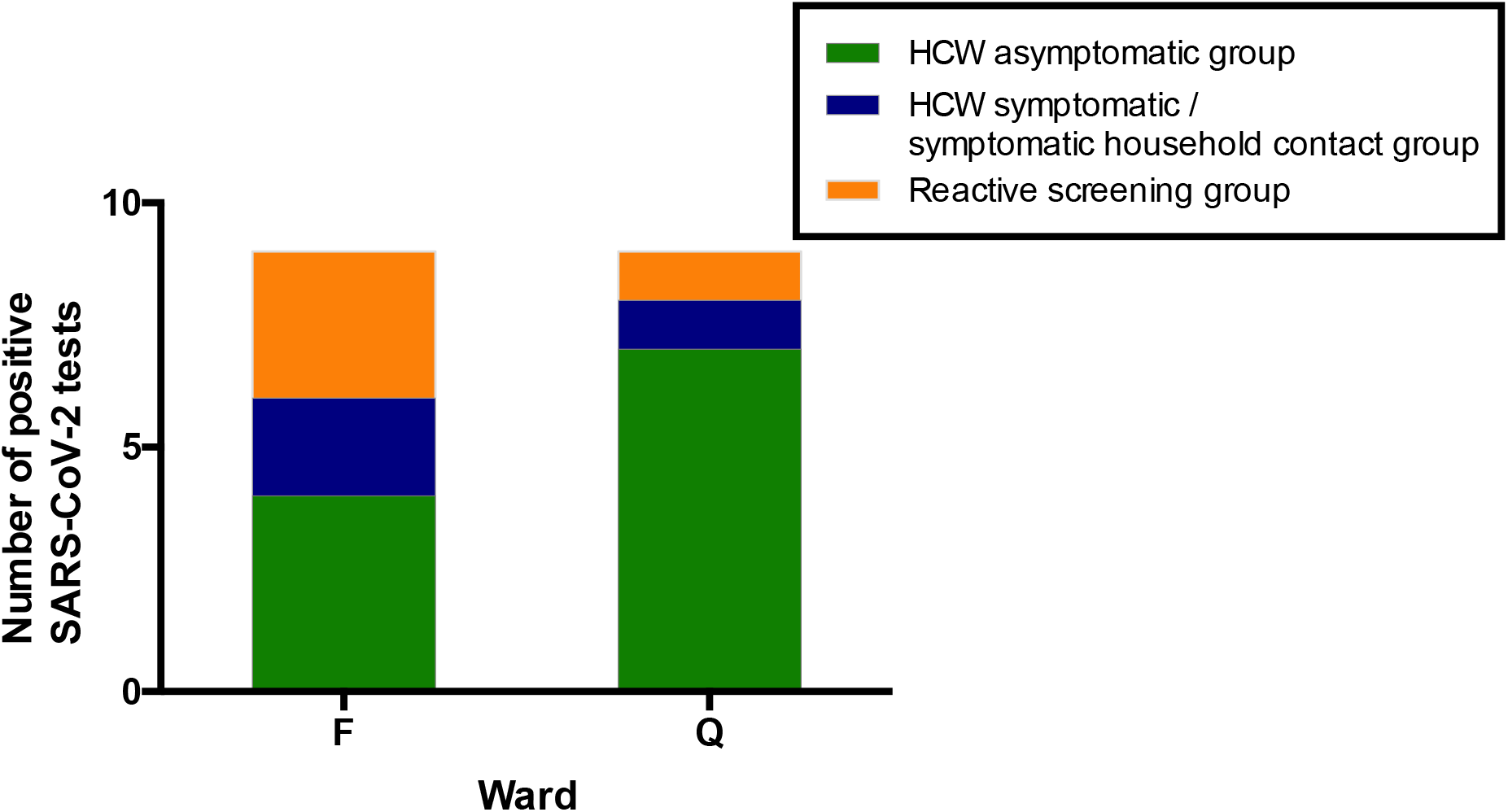
All SARS-CoV-2 positive HCW identified in Wards F and Q, stratified by their mechanism of identification. Individuals testing positive for SARS-CoV-2 in the ‘*HCW asymptomatic screening group* were identified by the asymptomatic screening programme. Individuals testing positive in the ‘*HCW symptomatic / symptomatic household contact groups’* were identified by self-presentation after developing symptoms. Individuals testing positive in the ‘*Reactive screening group* were identified by an intensified screening programme after initial positive clusters had been recognised.

Ward Q is a general medical ward designated as a ‘red’ clinical area for the care of COVID-19 positive patients, with a Scenario 1 PPE protocol (Tables S3-S4). Here, 7/37 (19%) ward staff tested positive for SARS-CoV-2. In addition, one staff member tested positive as part of the *HCW symptomatic screening group*, within the same period as ward surveillance. Reactive screening of a further five ward Q staff uncovered one additional infection. 4/4 sequenced viruses were of the B·1 lineage (Figures S2-S3, Table S6; other isolates could not be sequenced due to a sample CT value >30). All positive HCWs were requested to self-isolate, and infection control and PPE reviews were undertaken to ensure that environmental cleaning and PPE donning/doffing practices were compliant with hospital protocol. Staff training and education was provided to address observed instances of incorrect infection control or PPE practice.

Ward O, a ‘red’ medical ward, had similar numbers of asymptomatic HCWs screened as ward F, and a similar positivity rate (4/44; 9%). This ward was listed for further cluster investigation after the study ended, however incorrect PPE usage was not noted during the study period.

### Characteristics of the HCW symptomatic and HCW symptomatic-contact screening groups

The majority of individuals who tested positive for SARS-CoV-2 after screening due to the presence of symptoms had high COVID-19 probability (Table 3). This reflects national guidance regarding self-isolation at the time of our study.^22^

**Table 3.** Distribution of positive SARS-CoV-2 tests amongst symptomatic individuals with a positive test result, categorised according to test group and COVID-19 symptom-based probability criteria (as defined in Table S4).

| Distribution of COVID-19 clinical probability scores for individuals with a positive SARS-CoV-2 test result |  |  |  |  |
| --- | --- | --- | --- | --- |
|  | High | Medium | Low | Total |
| **HCW symptomatic screening group** | 22/26<br>(85%) | 3/26<br>(11%) | 1/26<br>(4%) | **26/26**<br>**(100%)** |
| **HCW symptomatic household contacts** | 3/4<br>(75%) | 0/4<br>(0%) | 1/4<br>(25%) | **4/4**<br>**(100%)** |

## Discussion

Through the rapid establishment of an expanded HCW SARS-CoV-2 screening programme, we discovered that 31/1,032 (3%) of HCWs tested positive for SARS-CoV-2 in the absence of symptoms. Of 30 individuals from this *asymptomatic screening group* studied in more depth, 6/30 (20%) had not experienced any symptoms at the time of their test. 1/6 became symptomatic suggesting that the true asymptomatic carriage rate was 5/1,032 (0.5%)). 11/30 (37%) had experienced mild symptoms prior to testing. Whilst temporally associated, it cannot be assumed that these symptoms necessarily resulted from COVID-19. These proportions are difficult to contextualise due to paucity of point-prevalence data from asymptomatic individuals in similar healthcare settings or the wider community. For contrast, 60% of asymptomatic residents in a recent study tested positive in the midst of a care home outbreak.^23^ Regardless of the proportion, however, many secondary and tertiary hospital-acquired infections were undoubtedly prevented by identifying and isolating these SARS-CoV-2 positive HCWs.

12/30 (40%) individuals from the *HCW asymptomatic screening group* reported symptoms >7 days prior to testing, and the majority experiencing symptoms consistent with a high probability of COVID-19 had appropriately self-isolated during that period. Patients with COVID-19 can remain SARS-CoV-2 PCR positive for a median of 20 days (IQR 17-24) after symptom onset^24^, and the limited data available suggest viable virus is not shed beyond eight days.^25^ A pragmatic approach was taken to allowing individuals to remain at work, where the HCW had experienced high probability symptoms starting >7 days and ≤1 month prior to their test and had been well for the preceding 48 hours. This approach was based on the following: low seasonal incidence of alternative viral causes of high COVID-19 probability symptoms in the UK^26^, the high potential for SARS-CoV-2 exposure during the pandemic and the potential for prolonged, non-infectious shedding of viral RNA.^24,25^ For other individuals, we applied standard national guidelines requiring isolation for seven days from the point of testing.^27^ However, for HCW developing symptoms after a positive swab, isolation was extended for seven days from symptom onset.

Our data clearly demonstrate that focusing solely on the testing of individuals fitting a strict clinical case definition for COVID-19 will inevitably miss asymptomatic and pauci-symptomatic disease. This is of particular importance in the presence of falling numbers of community COVID-19 cases, as hospitals will become potential epicentres of local outbreaks. Therefore, we suggest that in the setting of limited testing capacity, a high priority should be given to a reactive asymptomatic screening programme that responds in real-time to HCW sickness trends, or (to add precision) incidence of positive tests by area. The value of this approach is illustrated by our detection of a cluster of cases in ward F, where the potential for uncontrolled staff-to-staff or staff-to-patient transmission could have led to substantial morbidity and mortality in a particularly vulnerable patient group. As SARS-CoV-2 testing capacity increases, rolling programmes of serial screening for asymptomatic staff in all areas of the hospital is recommended, with the frequency of screening being dictated by anticipated probability of infection. The utility of this approach in care-homes and other essential institutions should also be explored, as should serial screening of long-term inpatients.

The early success of our programme relied upon substantial collaborative efforts between a diverse range of local stakeholders. Similar collaborations will likely play a key role in the rapid, *de novo* development of comprehensive screening programmes elsewhere. The full benefits of enhanced HCW screening are critically dependent upon rapid availability of results. A key success of our programme has been bespoke optimisation of sampling and laboratory workflows enabling same-day resulting, whilst minimising disruption to hospital processes by avoiding travel to off-site testing facilities. Rapid turnaround for testing and sequencing is vital in enabling timely response to localised infection clusters, as is the maintenance of reserve capacity to allow urgent, reactive investigations.

There appeared to be a significantly higher incidence of HCW infections in ‘red’ compared to ‘green’ wards. Many explanations for this observation exist, and this study cannot differentiate between them. Possible explanations include transmission between patients and HCW, HCW-to-HCW transmission, variability of staff exposure outside the workplace and non-random selection of wards. It is also possible that, even over the three weeks of the study, ‘red’ wards were sampled earlier during the evolution of the epidemic when transmission was greater. Further research into these findings is clearly needed on a larger scale. Furthermore, given the clear potential for pre-symptomatic and asymptomatic transmission amongst HCWs, and data suggesting that infectivity may peak prior to symptom onset^11^, there is a strong argument for basic PPE provision in all clinical areas.

The identification of transmission within the hospital through routine data is problematic. Hospitals are not closed systems and are subject to numerous external sources of infection. Coronaviruses generally have very low mutation rates (~10^-6^ per site per cycle)^28^, with the first reported sequence of the current pandemic only published on 12^th^ January 2020.^29^ In addition, given SARS CoV-2 was only introduced into the human population in late 2019, there is at present a lack of diversity in circulating strains. However, as the pandemic unfolds and detailed epidemiological and genome sequence data from patient and HCW clusters are generated, real-time study of transmission dynamics will become an increasingly important means of informing disease control responses and rapidly confirming (or refuting) hospital acquired infection. Importantly, implementation of such a programme would require active screening and rapid sequencing of positive cases in both the HCW and patient populations. Prospective epidemiological data will also inform whether hospital staff are more likely to be infected in the community or at work, and may identify risk factors for the acquisition of infection, such as congregation in communal staff areas or inadequate access to PPE.

Our study is limited by the relatively short time-frame, a small number of positive tests and a lack of behavioural data. In particular, the absence of detailed workplace and community epidemiological data makes it difficult to draw firm conclusions with regards to hospital transmission dynamics. The low rate of observed positive tests may be partly explained by low rates of infection in the East of England in comparison with other areas of the UK (cumulative incidence 0·17%, thus far).^30^ The long-term benefits of HCW screening on healthcare systems will be informed by sustained longitudinal sampling of staff in multiple locations. More comprehensive data will parametrise workforce depletion and COVID-19 transmission models. The incorporation of additional information including staffing levels, absenteeism, and changes in proportions of staff self-isolating before and after the introduction of widespread testing will better inform the impact of screening at a national and international level. Such models will be critical for optimising the impact on occupationally-acquired COVID-19, and reducing the likelihood that hospitals become hubs for sustained COVID-19 transmission.

In the absence of an efficacious vaccine, additional waves of COVID-19 are likely as social distancing rules are relaxed. Understanding how to limit hospital transmission will be vital in determining infection control policy, and retain its relevance when reliable serological testing becomes widely available. Our data suggest that the roll-out of screening programmes to include asymptomatic as well as symptomatic patient-facing staff should be a national and international priority. Our approach may also be of benefit in reducing transmission in other institutions, for example care-homes. Taken together, these measures will increase patient confidence and willingness to access healthcare services, benefiting both those with COVID-19 and non-COVID-19 disease.

## Materials and Methods

### Staff screening protocols

A full description of methods can be found in Supplementary Information. Briefly, two parallel streams of entry into the testing programme were established. The first was operated by the Occupational Health department and allowed any patient-facing or non-patient-facing hospital employee (HCW) to refer themselves or a household contact, including children, should they develop symptoms suggestive of COVID-19. The second was a rolling programme of testing for all patient-facing and non-patient-facing staff working in defined clinical areas thought to be at risk of SARS-CoV-2 transmission. A traffic-light colouring system was used to denote anticipated risk of SARS-CoV-2 transmission by ward (table S1), with different types of personal protective equipment (PPE) used in each (table S2). Inclusion into the programme was voluntary, and offered to all individuals working in a given ward during the time of sampling. Regardless of the route of entry into the programme, the process for testing and follow-up was identical. Wards were closed to external visitors.

Self-isolation and household quarantine advice was determined by estimating the pre-test probability of COVID-19 (high, medium or low) in those with symptoms, based on the presence or absence of typical features (tables S3-S4). Symptom history was obtained for all symptomatic HCWs at the time of self-referral, and again for all positive cases via telephone interview when results became available. All individuals who had no symptoms at the time of testing were followed up by telephone within 14 days of their result. Pauci-symptomatic individuals were defined as those with low-probability clinical COVID-19 criteria (table S3).

### Laboratory methods

The swabbing, extraction and amplification methods for this study follow a recently validated procedure.^16^ Individuals performed a self-swab at the back of the throat followed by the nasal cavity as previously described.^2^ The single dry sterile swab was immediately placed into transport medium/lysis buffer containing 4M guanidine thiocyanate to inactivate virus, and carrier RNA. This facilitated BSL2-based manual extraction of viral RNA in the presence of MS2 bacteriophage amplification control. Use of these reagents and components avoided the need for nationally employed testing kits. Real-time RT-PCR amplification was performed as previously described and results validated by confirmation of FAM amplification of the appropriate controls with threshold cycle (CT) ≤36. Lower CT values correspond to earlier detection of the viral RNA in the RT-PCR process, corresponding with a higher copy number of the viral genome. In 2/1,270 cases, RT-PCR failed to amplify the internal control and results were discarded, with HCW offered a re-test. Sequencing of positive samples was attempted on samples with a CT ≤30 using a multiplex PCR based approach^17^ using the modified ARTTC v2 protocol^18^ and v3 primer set.^19^ Genomes were assembled using reference based assembly and the bioinformatic pipeline as described^17^ using a 20x minimum coverage as a cut-off for any region of the genome and a 50·1% cut-off for calling of single nucleotide polymorphisms (SNPs). Samples were sequenced as part of the COVID-19 Genomics UK Consortium, COG-UK), a partnership of NHS organisations, academic institutions, UK public health agencies and the Wellcome Sanger Institute.

### Data extraction and analysis

Swab result data were extracted directly from the hospital-laboratory interface software, Epic (Verona, Wisconsin, USA). Details of symptoms recorded at the time of telephone consultation were extracted manually from review of Epic clinical records. Data were collated using Microsoft Excel, and figures produced with GraphPad Prism (GraphPad Software, La Jolla, California, USA). Fisher’s exact test was used for comparison of positive rates between groups defined in the main text. Mann-Whitney testing was used to compare CT values between different categories of tested individuals. HCW samples that gave SARS CoV-2 genomes were assigned global lineages defined by ^20^ using the PANGOLIN utility.^21^

### Ethics and consent

As a study of healthcare-associated infections, this investigation is exempt from requiring ethical approval under Section 251 of the NHS Act 2006 (see also the NHS Health Research Authority algorithm, available at http://www.hradecisiontools.org.uk/research/, which concludes that no formal ethical approval is required). Written consent was obtained from each HCW described in the anonymised case vignettes.

### Funding

No funding sources have had any role in data collection, analysis, interpretation, writing of the manuscript or the decision to submit for publication. No authors have been paid to write this article by a pharmaceutical company or any other agency. LR, DS, MR, NKJ, IG, SB, MPW had access to all the data. IG, SB, MPW held final responsibility for the decision to submit the manuscript for publication.

## Data Availability

Sequencing data have been deposited in GSAID under accession codes EPI_ISL_433989 - EPI_ISL_433992, EPI_ISL_434005, EPI_ISL_433489 -
EPI_ISL_433497. The authors confirm all other data supporting the findings of this study are available within the article and its supplementary materials.

## Acknowledgements

This work was supported by the Wellcome Trust Senior Research Fellowships 108070/Z/15/Z to MPW, 215515/Z/19/Z to SGB and 207498/Z/17/Z to IGG; Collaborative award 206298/B/17/Z to IGG; Principal Research Fellowship 210688/Z/18/Z to PJL; Investigator Award 200871/Z/16/Z to KGCS; Addenbrooke’s Charitable Trust (to MPW, SGB, IGG and PJL); the Medical Research Council (CSF MR/P008801/1 to NJM); NHS Blood and Transfusion (WPA15-02 to NJM); National Institute for Health Research (Cambridge Biomedical Research Centre at CUHNFT), to JRB, MET, AC and GD, Academy of Medical Sciences and the Health Foundation (Clinician Scientist Fellowship to MET), Engineering and Physical Sciences Research Council (EP/P031447/1 and EP/N031938/1 to RS),Cancer Research UK (PRECISION Grand Challenge C38317/A24043 award to JY). Components of this work were supported by the COVID-19 Genomics UK Consortium, (COG-UK), which is supported by funding from the Medical Research Council (MRC) part of UK Research & Innovation (UKRI), the National Institute of Health Research (NIHR) and Genome Research Limited, operating as the Wellcome Sanger Institute

## Conflict of Merest statements

Lucy Rivett has nothing to disclose.

Sushmita Sridhar has nothing to disclose.

Dominic Sparkes has nothing to disclose.

Matthew Routledge has nothing to disclose.

Nick Jones has nothing to disclose.

Sally Forrest has nothing to disclose.

Jamie Young has nothing to disclose.

Joana Pereira-Dias has nothing to disclose.

William Hamilton has nothing to disclose.

Mark Ferris has nothing to disclose.

M. Estee Torok reports grants from Academy of Medical Sciences and the Health Foundation, non-financial support from National Institute of Health Research, during the conduct of the study; grants from Medical Research Council, grants from Global Challenges Research Fund, personal fees from Wellcome Sanger Institute, personal fees from University of Cambridge, personal fees from Oxford University Press, outside the submitted work.

Luke Meredith has nothing to disclose.

Neil Bartholomew has nothing to disclose.

Surendra Parmar has nothing to disclose.

Martin Curran has nothing to disclose.

Stewart Fuller has nothing to disclose.

Afzal Chaudhry reports grants from Cambridge Biomedical Research Centre at CUHNFT, during the conduct of the study.

Ashley Shaw has nothing to disclose.

Richard J. Samsworth reports grants from EPSRC fellowship, during the conduct of the study.

John Bradley has nothing to disclose.

Gordon Dougan reports grants from NIHR, during the conduct of the study.

Kenneth G.C. Smith reports grants from Wellcome Trust, during the conduct of the study.

Paul J. Lehner reports grants from Wellcome Trust Principal Research Fellowship, grants from Addenbrooke’s Charitable Trust, during the conduct of the study.

Nicholas J. Matheson reports grants from Medical Research Council (Clinician Scientist Fellowship), grants from NHS Blood and Transfusion, during the conduct of the study.

Giles Wright has nothing to disclose.

Ian Goodfellow reports grants from Wellcome Trust (Senior Research Fellowships), grants from Wellcome Trust (Collaborative Award), grants from Addenbrooke’s Charitable Trust, during the conduct of the study.

Stephen Baker reports grants from Wellcome Trust (Senior Research Fellowships), from Addenbrooke’s Charitable Trust, during the conduct of the study.

Michael P. Weekes reports grants from Wellcome Trust (Senior Research Fellowships), from Addenbrooke’s Charitable Trust, during the conduct of the study.

## Supplementary Material

### Further details of HCW screening protocols

The *HCW symptomatic* and *HCW household contact* screening programme was managed jointly by the Occupational Health and Infectious Diseases departments. For the *HCW asymptomatic group*, the hospital’s system for categorising clinical areas according to COVID-19 risk is summarised in Table S1. Daily workforce sickness reports and trends in the results of HCW testing were monitored to enable areas of concern to be highlighted and targeted for screening and cluster analysis, in a reactive approach. High throughput clinical areas where staff might be exposed to large numbers of suspected COVID-19 patients were also prioritised for staff screening. These included the Emergency Department, the COVID-19 Assessment Unit, and a number of ‘red’ inpatient wards. Staff caring for the highest priory ‘shielding’ patients (Haematology/Oncology, Transplant medicine) were also screened, as were a representative sample of staff from ‘Amber’ and ‘Green’ areas. The personal protective equipment (PPE) worn by staff in these areas is summarised in Table S2.

### Occupational Health Triage and Appointment Booking

We devised a scoring system to determine the clinical probability of COVID-19 based on symptoms from existing literature^1,2^ (Table S3). Self-referring HCW and staff captured by daily workforce sickness reports were triaged by designated Occupational Health nurses using these criteria (Table S4). Self-isolating staff in the medium and low probability categories were prioritised for testing, since a change in the clinical management was most likely to derive from results.

### Sample collection procedures

Testing was primarily undertaken at temporary on-site facilities. Two ‘Pods’ (self-contained portable cabins with office, kitchen facilities, generator and toilet) were erected in close proximity both to the laboratory and main hospital. Outside space was designed to enable car and pedestrian access, and ensure ≥2 m social distancing at all times. Individuals attending on foot were given pre-prepared self-swabbing kits containing a swab, electronically labelled specimen tube, gloves and swabbing instructions contained in a zip-locked collection bag. Pods were staffed by a team of re-deployed research nurses, who facilitated self-swabbing by providing instruction as required. Scenario 1 PPE (Table S2) was worn by Pod nurses at all times. Individuals in cars were handed self-swabbing kits through the window, with samples dropped in collection bags into collection bins outside. Any children (household contacts) were brought to the pods in cars and swabbed in situ by a parent or guardian.

In addition to Pod-based testing, an outreach *HCW asymptomatic screening* service was developed to enable self-swabbing kits to be delivered to HCWs in their area of work, minimising disruption to the working routine of hospital staff, and maximising Pod availability for symptomatic staff. Lists of all staff working in target areas over a 24-hour period were assembled, and kits pre-prepared accordingly. Self-swabbing kits were delivered to target areas by research nurses, who trained senior nurses in the area to instruct other colleagues on safe self-swabbing technique. Kits were left in target areas for 24 hours to capture a full cycle of shift patterns, and all kits and delivery equipment were thoroughly decontaminated with 70% ethanol prior to collection. Twice daily, specimens were delivered to the laboratory for processing.

### Results reporting

As soon as they were available, positive results were telephoned to patients by Infectious Diseases physicians, who took further details of symptomatology including timing of onset, and gave clinical advice (Table S4). Negative results were reported by Occupational Health nurses via telephone, or emailed through a secure internal email system. Advice on returning to work was given as described in Table S4. Individuals advised to self-isolate were instructed to do so in their usual place of residence. Particularly vulnerable staff or those who had more severe illness but did not require hospitalisation were offered follow-up telephone consultations. Individuals without symptoms at the time of testing were similarly followed up, to monitor for *de novo* symptoms. Verbal consent was gained for all results to be reported to the hospital’s infection control and health & safety teams, and to Public Health England, who received all positive and negative results as part of a daily reporting stream.

**Table S1.** Categories of ward according to anticipated COVID-19 exposure risk.

| **Red (high risk)** | **Amber (medium risk)** | **Green (low risk)** |
| --- | --- | --- |
| Areas with confirmed SARS-CoV-2 RT-PCR positive patients, or patients with very high clinical suspicion of COVID-19 | Areas with patients awaiting SARS-CoV-2 RT-PCR test results, or that have been exposed and may be incubating infection | Areas with no known SARS-CoV-2 RT-PCR positive patients, and none with clinically suspected COVID-19 |

**Table S2.** Categories of clinical area according to personal protective equipment (PPE) protocol (‘PPE Scenarios’)

| **Personal protective equipment (PPE) 'Scenarios'** |  |  |  |  |
| --- | --- | --- | --- | --- |
|  | **Scenario 0** | **Scenario 1** | **Scenario 2** | **Scenario 3** |
| **Area description** | All clinical areas without any known or suspected COVID-19 cases | Designated ward, triage and assessment-based care with suspected or confirmed COVID-19 patients | Cohorted areas where aerosol-generating procedures are carried out frequently with suspected or confirmed COVID-19 patients | Operating theatres where procedures are performed with suspected or confirmed COVID-19 patients |
| **PPE description** | Fluid resistant face mask at all times, apron and non-sterile gloves for patient contact (within two metres) | Surgical scrubs, fluid resistant face mask, theatre cap, eye protection, apron and non-sterile gloves | Water repellent gown, FFP3 mask, eye protection, theatre cap, surgical gloves, with an apron and non-sterile gloves in addition for patient contact (within two metres) | Water repellent gown, FFP3 mask, eye protection, theatre cap and surgical gloves |
| **Ward categories** | Green wards, e.g. designated areas of emergency department and medical admissions unit. Medical, surgical and haematology wards / outpatient clinics. | Amber + red wards, e.g. designated areas of emergency department and medical admissions unit. Designated CoVID-19 confirmed wards. | Amber + red wards, e.g. intensive care unit, respiratory units with non-invasive ventilation facilities. | All operating theatres, including facilities for bronchoscopy and endoscopy. |

All users of FFP3 masks underwent routine fit-testing prior to usage. Cleaning and re-use of masks, theatre caps, gloves, aprons or gowns was actively discouraged. Cleaning and re-use of eye protection was permitted for certain types of goggles and visors, as specified in the hospital’s PPE protocol. Single-use eye protection was in use in most Scenario 1 and 2 areas, and was not cleaned and re-used. All non-invasive ventilation or use of high-flow nasal oxygen on laboratory-confirmed or clinically suspected COVID-19 patients was performed in negative-pressure (−5 pascals) side rooms, with 10 air changes per hour and use of Scenario 2 PPE. All other aerosol generating procedures were undertaken with Scenario 2 PPE precautions, in negative- or neutral-pressure facilities. General clinical areas underwent a minimum of 6 air changes per hour, but all critical care areas underwent a minimum of 10 air changes per hour as a matter of routine. Surgical operating theatres routinely underwent a minimum of 25 air changes per hour.

**Table S3.** Clinical criteria for estimating pre-test probability of COVID-19.

| **COVID-19 probability criteria** |  |
| --- | --- |
| Major | Fever ($>37.8^{\circ}\text{C}$) |
|  | New persistent cough |
|  | Unprotected close contact with a confirmed case* |
| Minor | Hoarse voice |
|  | Non-persistent cough |
|  | Sore throat |
|  | Nasal discharge or congestion |
|  | Shortness of breath |
|  | Wheeze |
|  | Headache |
|  | Muscle aches |
|  | Nausea and/or vomiting and/or diarrhoea |
|  | Loss of sense of taste or smell |
| *Unprotected close contact defined as either face-to-face contact or spending more than 15 minutes within 2 metres of an infected person, without wearing appropriate PPE. |  |

**Table S4.** Categories of pre-test probability of COVID-19, according to the presence of clinical features shown in table S1.

| Stratification of COVID-19 probability |  | Implications for exclusion from work |
| --- | --- | --- |
| High probability | $\geq 2$ major symptoms<br>or<br>$\geq 1$ major symptom and<br>$\geq 2$ minor symptoms | Self-isolate for 7 days from the date of onset, regardless of the test result. Only return to work if afebrile for 48 hours and symptoms have improved*.<br>Household contacts should self-quarantine for 14 days from the date of symptom onset in the index case, regardless of the test result. If they develop symptoms, they should self-isolate for 7 days from the date of onset, and only return to work if afebrile for 48 hours and symptoms have improved*. |
| Medium probability | 1 major symptom<br>or<br>0 major symptoms and<br>$\geq 3$ minor symptoms | Test result positive: self-isolate for 7 days from the date of onset, and only return to work if afebrile for 48 hours and symptoms have improved*. Household contacts should self-quarantine for 14 days from the date of index case symptom onset. If they develop symptoms, they should self-isolate for 7 days from the date of onset, and only return to work if afebrile for 48 hours and symptoms have improved*.<br>Test result negative: repeat testing at 48 hours from the initial swab. If repeat testing is positive, follow the advice detailed above. If repeat testing is negative, return to work, unless symptoms worsen. Self-quarantine not required for household contacts. |
| Low probability | 0 major symptoms and 1-2 minor symptoms | Test result positive: self-isolate for 7 days from the date of test, and only return to work if afebrile for 48 hours and symptoms have improved*. Household contacts should self-quarantine for 14 days from the date of test. If they develop symptoms, they should self-isolate for 7 days from the date of onset, and only return to work if afebrile for 48 hours and symptoms have improved*.<br>Test result negative: return to work, unless symptoms worsen. Self-quarantine not required for household contacts. |
| Asymptomatic | 0 major symptoms and 0 minor symptoms | Test result positive: self-isolate for 7 days from the date of test. If symptoms develop after the test, self-isolation should occur for 7 days from the date of onset, and return to work should only occur if afebrile for 48 hours and symptoms have improved*. Household contacts should self-quarantine for 14 days from the date of the test. If they develop symptoms, they should self-isolate for 7 days from the date of onset, and only return to work if afebrile for 48 hours and symptoms have improved*.<br>Test result negative: continue working, unless symptoms develop. Self-quarantine not required for household contacts. |

\*Residual cough in the absence of other symptoms should not preclude returning to work.

**Table S5:** Details of distribution of SARS-CoV-2 positive cases across 21 clinical areas selected for systematic ward-based asymptomatic screening (shown as Figure 3 in the main text) Areas are coloured (‘green’, ‘amber’, ‘red’) according to anticipated risk of exposure to COVID-19 (Table S1). HCWs working across >1 ward were counted for each area. In addition a number of individuals from other clinical areas were tested on an opportunistic basis, none of whom tested positive (not shown here but included in HCW asymptomatic screening group in Table 1), and an enhanced screening programme was subsequently provided for HCWs on wards F and Q (Figure 4).

|  | Clinical area / ward | Positive tests / Total tests |
| --- | --- | --- |
| Green | A | 0/26 |
|  | B | 1/44 |
|  | C | 0/30 |
|  | D | 0/38 |
|  | E | 1/54 |
|  | F | 4/43 |
|  | G | 0/15 |
|  | H | 0/12 |
|  | I | 0/17 |
|  | J | 0/31 |
| Amber | K | 3/54 |
| Red | L | 0/11 |
|  | M | 1/35 |
|  | N | 0/20 |
|  | O | 4/44 |
|  | P | 0/19 |
|  | Q | 7/37 |
|  | R | 3/72 |
|  | S | 1/59 |
|  | T | 2/42 |
|  | U | 1/33 |
| All areas |  | 28/736 |

**Table S6:** Details of each SARS-CoV-2 positive isolate from all HCWs and household contacts in the study.

| Patient ID | Type | HCW_ward | Ct value | Seq Attempted | Seq_ID | % Sequence Coverage | Average Seq Depth | PANGOLIN lineage |
| --- | --- | --- | --- | --- | --- | --- | --- | --- |
| C1 | Symptomatic Contact | HCW Contact | 23.9 | Y | CAMB-7FBB0 | 99.61 | 2048.5 | B.1 |
| C3 | Symptomatic Contact | HCW Contact | 23 | N |  |  |  | Not available |
| H3 | Asymptomatic | B | 31 | Y | CAMB-7C0C3 | 98.61 | 835.084 | B.1 |
| H54 | Symptomatic | B | 35 |  |  |  |  |  |
| H12 | Symptomatic | C | 16 | Y | CAMB-7FB92 | 99.60 | 3312.22 | B.1 |
| H19 | Asymptomatic | E | 27 | Y | CAMB-7FC26 | 99.61 | 3632.26 | B.1.1 |
| C2 | Asymptomatic | F | 15.5 | Y | CAMB-7FC08 | 99.60 | 3157.08 | B.1 |
| H17 | Asymptomatic | F | 33.6 | Y | CAMB-7FBFC | 99.61 | 1167.76 | B.1 |
| H20 | Asymptomatic | F | 18 | Y | CAMB-7FC35 | 99.61 | 1350.65 | B.1 |
| H21 | Asymptomatic | F | 22.8 | Y | CAMB-7FC44 | 99.60 | 3584.79 | B.1 |
| H22 | Symptomatic | F | 24 | Y | CAMB-7FC53 | 99.60 | 3692.14 | B.1.7 |
| H23 | Asymptomatic | F | 32.7 | Y | CAMB-7FC62 | 99.60 | 1610.33 | B.2.1 |
| H35 | Symptomatic | F | 36 | Y | CAMB-8221F | 73.00 | 104.391 | B.1 |
| H36 | Asymptomatic | F | 29 | Y | CAMB-8222E | 98.59 | 1882.65 | B.1.7 |
| H53 | Symptomatic Contact | HCW Contact | 23 |  |  |  |  |  |
| H38 | Asymptomatic | K | 36 | N |  |  |  | Not available |
| H39 | Asymptomatic | K | 31 | N |  |  |  | Not available |
| H28 | Symptomatic | K/R/L/T/OTHER | 18 | Y | CAMB-7FD32 | 99.60 | 3770.36 | B.1.11 |
| H11 | Asymptomatic | M | 32 | Y | CAMB-7FB83 | 99.60 | 1044.43 | B.1 |
| H32 | Symptomatic | N | 33 | Y | CAMB-81007 | 97.62 | 1196.53 | B.1 |
| H47 | Symptomatic | N | 32 | N |  |  |  | Not available |
| H31 | Asymptomatic | O | 29 | Y | CAMB-80FFC | 99.59 | 2286.08 | B.1 |
| H45 | Asymptomatic | O | 36 | N |  |  |  | Not available |
| H51 | Symptomatic | O | 33 |  |  |  |  |  |
| H57 | Symptomatic Contact | O | 23 |  |  |  |  |  |
| H1 | Asymptomatic | OTHER | 23 | Y | CAMB-7C0A5 | 98.61 | 2277.92 | B.1 |
| H6 | Symptomatic | OTHER | 30 | Y | CAMB-7FB29 | 98.75 | 1317.43 | B.1 |
| H7 | Symptomatic | OTHER | 26 | Y | CAMB-7FB47 | 99.61 | 3599.59 | B.1 |
| H10 | Symptomatic | OTHER | 22 | Y | CAMB-7FB74 | 99.60 | 187.059 | B.1 |
| H14 | Symptomatic | OTHER | 34 | Y | CAMB-7FBCF | 99.61 | 1066.74 | B.1 |
| H16 | Asymptomatic | OTHER | 27.8 | Y | CAMB-7FBED | 99.60 | 796.874 | B.1 |
| H24 | Symptomatic | OTHER | 21 | Y | CAMB-7FC80 | 98.62 | 916.884 | B.1 |
| H25 | Symptomatic | OTHER | 21 | Y | CAMB-7FC9F | 99.60 | 1505.09 | B.1 |
| H33 | Symptomatic | OTHER | 35 | Y | CAMB-81016 | 90.92 | 233.779 | B.1 |
| H40 | Symptomatic | OTHER | 23 | N |  |  |  | Not available |
| H46 | Asymptomatic | OTHER | 36 | N |  |  |  | Not available |
| H55 | Symptomatic | Other | 26 |  |  |  |  |  |
| H56 | Asymptomatic | Other | 32 |  |  |  |  |  |
| H30 | Asymptomatic | OTHER/K/O/F | 31 | Y | CAMB-80FDE | 98.61 | 1773.74 | B.1 |
| H5 | Symptomatic | Q | 24 | Y | CAMB-7C1A2 | 97.75 | 2342.24 | B.1 |
| H8 | Symptomatic | Q | 14 | Y | CAMB-7FB56 | 99.60 | 2452.25 | B.1 |
| H18 | Asymptomatic | Q | 30 | Y | CAMB-7FC17 | 99.60 | 2585.89 | B.1 |
| H29 | Asymptomatic | Q | 31 | Y | CAMB-80AFB | 99.60 | 2028.31 | B.1 |
| H42 | Asymptomatic | Q | 35 | N |  |  |  | Not available |
| H44 | Asymptomatic | Q | 28 | N |  |  |  | Not available |
| H48 | Asymptomatic | Q | 36 | N |  |  |  | Not available |
| H49 | Asymptomatic | Q | 35 | N |  |  |  | Not available |
| H4 | Symptomatic | R | 24 | Y | CAMB-7C0D2 | 98.74 | 2083.89 | B.1 |
| H9 | Symptomatic | R | 19 | Y | CAMB-7FB65 | 99.61 | 3288.11 | B.1 |
| H13 | Symptomatic | R | 21 | Y | CAMB-7FBA1 | 99.60 | 3307.61 | B.1 |
| H27 | Asymptomatic | R | 25 | Y | CAMB-7FCBD | 98.61 | 1085.78 | B.1 |
| H34 | Symptomatic | R | 30 | Y | CAMB-81025 | 99.60 | 1997.98 | B.1 |
| H37 | Asymptomatic | R | 35 | N |  |  |  | Not available |
| H52 | Asymptomatic | R | 34 |  |  |  |  |  |
| H58 | Symptomatic | R/S/A/Q/P/L/N/M/K/Other | 24 |  |  |  |  |  |
| H15 | Symptomatic | S/N | 32 | Y | CAMB-7FBDE | 99.60 | 2246.43 | B.1.7 |
| H41 | Asymptomatic | S/Q | 31 | N |  |  |  | Not available |
| H2 | Asymptomatic | T | 36 | Y | CAMB-7C0B4 | 93.55 | 293.223 | B.1 |
| H26 | Asymptomatic | T | 32 | Y | CAMB-7FCAE | 0.03 | 0.189437 | Not available |
| H50 | Symptomatic | T | 34 | N |  |  |  | Not available |
| H43 | Asymptomatic | U | 32 | N |  |  |  | Not available |

**Figure S1:**
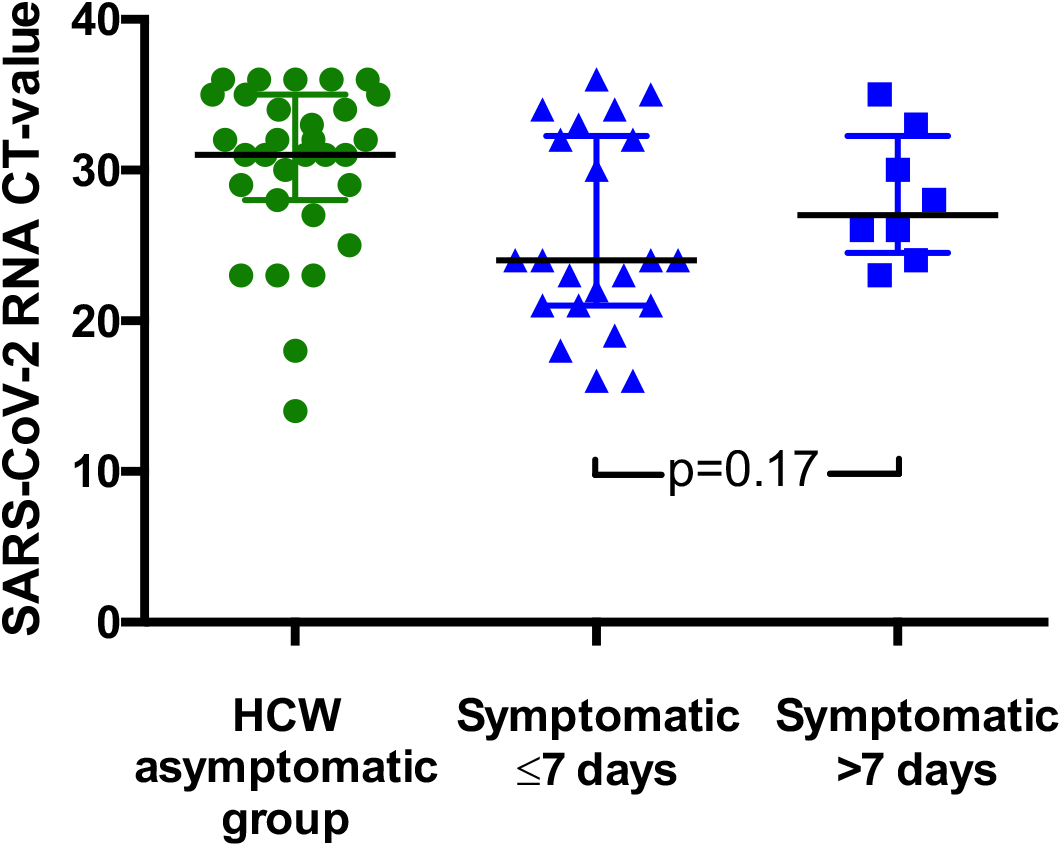
SARS-CoV-2 RNA CT values for HCWs testing positive according to presence and duration of symptoms. Results from the HCW symptomatic and HCW symptomatic contact groups are considered together in this figure. Individual CT values are shown, along with median and interquartile range for each group.

**Figure S2.**
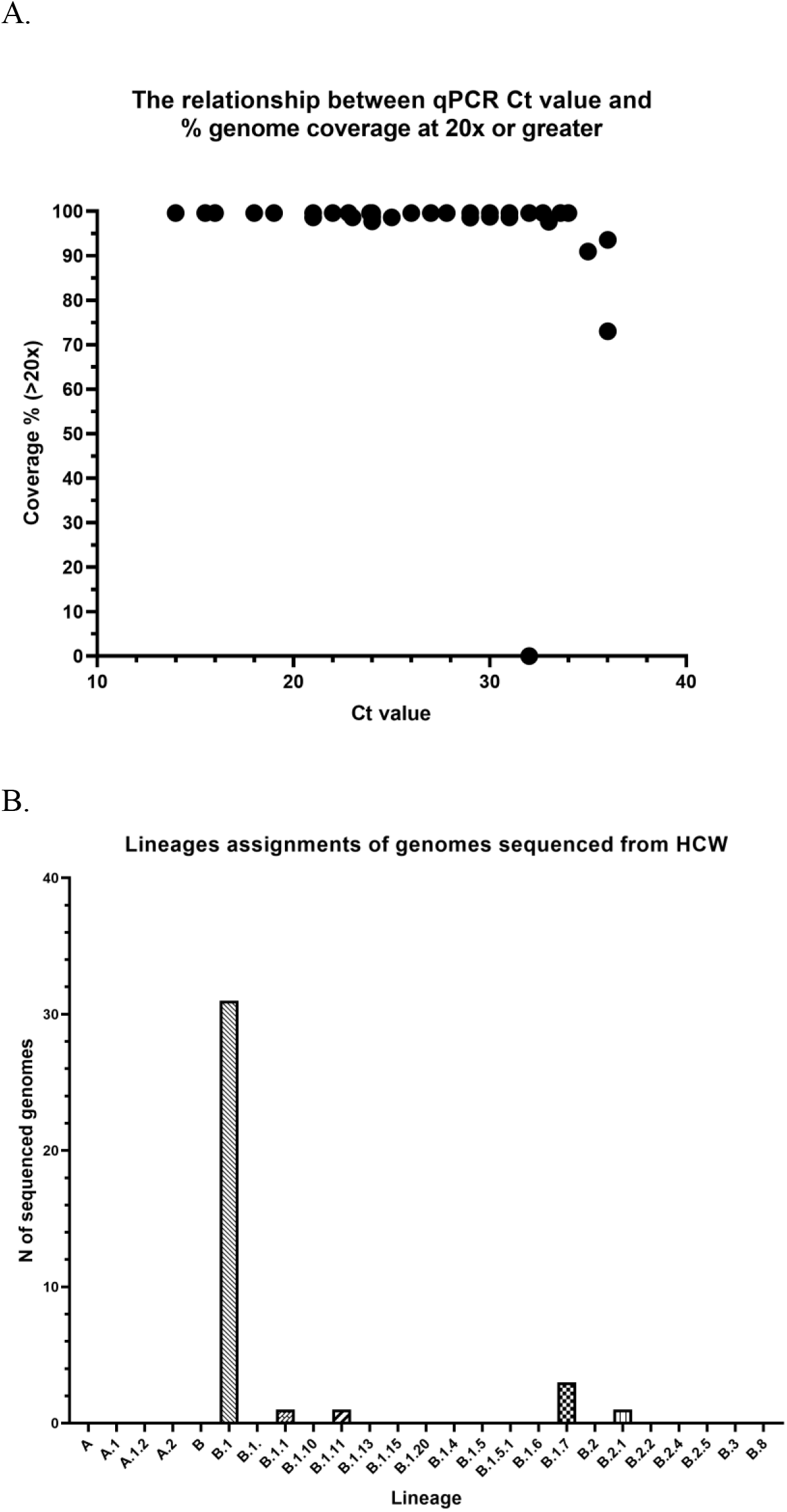
Further details of sequencing data. A) Comparisons of sequencing success rate vs Ct of HCW samples. Samples with CT less than 33 typically yielded genomes >90% coverage at a minimum depth of 20x. B) Lineage assignment of SARS CoV-2 genomes from HCW positive samples. Lineage assignments were generated using the PANGOLIN utility using a comparison against all currently circulating reference lineages.

**Figure S3.**
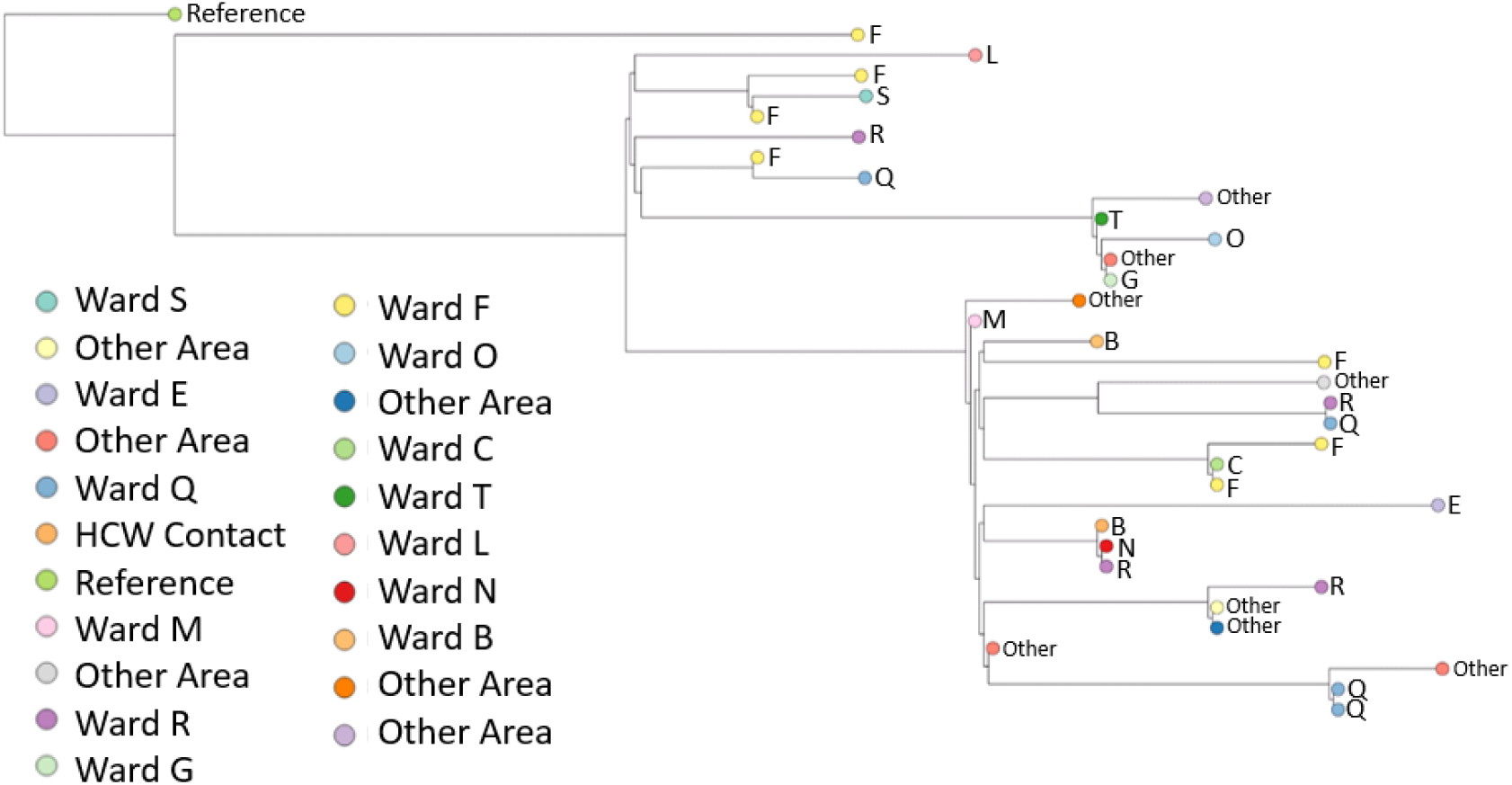
Phylogenetic tree of 34 healthcare worker (HCW) SARS-CoV-2 genomes. Branch tips are coloured by HCW base ward. 34/35 sequenced genomes passed the filter of <2990 (~10%) N. A SARS-CoV-2 genome collected in Wuhan in December 2019 was selected to root the tree, visualised initially on Nextstrain (https://nextstrain.org/) and the fasta file was downloaded from GISAID (ID: EPI ISL 402123) (https://www.gisaid.org/). Multiple sequence alignment of consensus fasta files was performed using MAFFT^3^ with default settings. The alignment was manually inspected using AliView.^4^ A maximum likelihood tree was produced using IQ-TREE^5^ software with ModelFinder Plus option (-m MFP), which chooses the nucleotide substitution model that minimises Bayesian information criterion (BIC) score. The model “chosen” was TPM2u+F (details: http://www.iqtree.org/doc/Substitution-Models). The tree was manually inspected in FigTree^6^, rooted on the 2019 Wuhan sample, ordered by descending node and exported as a Newick file. The tree was visualised in the online software Microreact^7^ in a private account, exported as a png image, which is shown here. Due to low genetic diversity in the virus (very recent introduction) genomic similarity alone cannot be used to infer transmission chains, as viruses can be identical by chance. Achieving higher resolution on transmission chains requires integrating clinical and detailed epidemiological data with genomic data from HCW and patients to uncover plausible transmission pathways.

